# The Association Between Tobacco Exposure During Pregnancy and Newborns’ Birth Weight in DKI Jakarta Community Members

**DOI:** 10.1101/2020.10.29.20222059

**Authors:** Jason Phowira, Felicitas Tania Elvina, Igor Ian Wiguna, Fathurohman Ramadhan Hanif Bari Wahyudi, Bernie Endyarni Medise

## Abstract

Low birth weight (LBW), a major determinant of neonate morbidity and mortality, remains a global public health concern. Intrauterine exposure to tobacco has been discerned as an important risk factor for LBW. This study aims to investigate the association between parental smoking during pregnancy and LBW. An analytical cross-sectional study was conducted from December 2019 - July 2020 on a random sample of parents with child aged 0-5 years old from 5 health centers in DKI Jakarta, Indonesia. A total of 145 subjects met the criteria and were analysed. Data analysis was carried out using IBM SPSS Statistics software. In the study, 11% of infants were born with LBW. The prevalence of smoking in fathers and mothers were 55.2% and 3.4%, respectively. Paternal smoking status was significantly associated with LBW (p < 0.05). Although not statistically significant, there was a dose-response relationship between paternal number of cigarettes/day and duration of smoking with LBW. Maternal smoking status (p = 0.448) was not closely associated with LBW, which might be due limited number of actively smoking mothers. From multivariate logistic regression, paternal smoking status, premature delivery, birth order and inadequate food intake during pregnancy were significant predictors of LBW (p < 0.05).

## Introduction

Smoking remains as the leading preventable cause of health problems and premature deaths globally. Despite acknowledging the harm and detrimental health effects, majority of highly dependent smokers find it arduous to quit smoking. The underlying reason behind this is primarily due to nicotine in cigarette generating strong desire to smoke, therefore overwhelming the concerns regarding its health impacts.^1^

Without the implementation of reduction strategies, tobacco use is projected over the next few decades to be the most significant cause of adult deaths worldwide.^2^ Indonesia is one of the countries with the most tobacco-consuming population, and the prevalence of tobacco users in Indonesia has been showing an increasing trend. In general, people in Indonesia begin casually smoking at an early age between 15 to 20, reflecting the vulnerability to initiate an early smoking habit among Indonesian youths.^3^ Tobacco is a strong and powerful multisite carcinogen with a global health impact, resulting in cancers of lungs, upper aero-digestive tracts, pancreas, liver, stomach, lower urinary tracts, kidney, and leukimia.^4^ About half of tobacco users die due to a smoking-related disease, and smoking reduces greatly one’s overall life expectancy by an average of 15 to 20 years.^5^

When associated with pregnancy, tobacco exposure can result in unfavorable pregnancy outcomes, including low birth weight (LBW), spontaneous abortion, premature delivery, perinatal mortality, and ectopic pregnancy.^6^ It has been long proposed and widely-believed that many of the chemicals present in tobacco can cross the placental barrier and lead to a direct harmful effect on the fetus.^7^ Organ volume analysis on magnetic resonance imaging (MRI) also demonstrated that *in utero* exposure to nicotine results in subsequent decreased brain, lung, kidney, and placental volume.^8^ The severity of fetal growth restriction (FGR) resulted from nicotine exposure has been demonstrated to be dose-dependent. Each pack smoked during pregnancy results in an average of 2.8 g decrement in neonatal body weight.^9^

Intrauterine growth restriction (IUGR) and low birth weight are notable risk factors of neonatal mortality. In United States, 65% of infant deaths occur among low birth weight infants, and in Indonesia, it was found that infants born with low birth weight had a 9.89-fold higher risk of neonatal death compared with infants born with normal birth weight.^10,11^ Neonatal mortality rate (NMR) remains high in Indonesia, with the rate of 21.4 infant deaths per 1,000 live births.^12^

Recent years have seen the emergence of several studies addressing the association between exposure to tobacco and newborns’ birth weight in other countries. However, to our knowledge, limited research has been dedicated on this topic in Indonesia. Thus, this research is conducted to analyze the association between paternal tobacco exposure during pregnancy and its effects on the infants’ birth weight at population level in Indonesia, particularly in DKI Jakarta. Hopes are the findings of this research could help provide better understandings about the harmful effects of tobacco exposure during pregnancy, leading to increased awareness and reduction in the number of morbidity and mortality associated with smoking.

## Methods

On this research, cross-sectional study was used to analyze the association between exposure to tobacco during pregnancy and birth weight of newborns. This research was performed for 8 months starting from December 2019 until July 2020, and the data collection was conducted at Primary Health Centers in DKI Jakarta, particularly in Central Jakarta. The target population were parents with child aged 0-5 years old from DKI Jakarta community members. The sample were the target population who fulfilled the inclusion and exclusion criteria. The inclusion criteria includes subjects who are being domiciled in DKI Jakarta, subjects with child aged 0-5 years possessing *Kesehatan Ibu dan Anak* (KIA) book or medical record from Primary Health Center, and subjects who agree to sign the informed consent. The exclusion criteria includes subjects with no data of birth weight written in KIA book or medical record, subjects with adopted child and subjects who do not fill the questionnaire completely.

Table 2×2 whose data was taken from previous study regarding the association between exposure to tobacco and low birth weight is used to obtain the sample size. The minimum sample required for this study was 98 subjects. In this research, the samples were selected based on simple random sampling method. The data was obtained from the questionnaires filled by eligible subjects. The questionnaire was modified from previous studies discussing about the association between tobacco exposure and several other factors related to prevalence of LBW. The data was used to analyze the association between tobacco exposure during pregnancy and the birth weight of the child in DKI Jakarta community members. In this research, the process of data collection was done by distributing printed questionnaires to obtain primary data regarding the exposure to tobacco during smoking and birth weight of the child.

Firstly, formal requests and ethical clearance were sent out to Primary Health Centers located in DKI Jakarta to ask for their permission. Then, randomized and eligible subjects were given explanation regarding the procedure and aim of the research. Subjects would have to read the informed consent form and asked to sign it if they agree with the conditions. Subjects were made aware of their rights in withdrawing from the research without any consequence and of their rights in relation to confidentiality. Then, the subjects filled the demographic data of the respondent followed by the questionnaire form. The researcher did not interfere with the subjects’ answers and helped the subjects in answering if there is any difficulty.

Data from this research was classified into two categories, low birth weight and normal birth weight. Low birth weight is defined as birth weight of less than 2500 grams, while ≥ 2500 grams is considered normal. All the data were analysed with the help of computer program SPSS version 20. The data were analysed as bivariate analysis using chi-square method, with the result being interpreted as significant if the p value is lower than 0.05. Variables showing p value of lower than 0.25 were analysed in multivariate logistic regression analysis to identify which variables were statistically significant after interrelations between different variables are controlled. The variable in logistic regression was considered significant if the p value is lower than 0.05.

## Results

### Study Population

The data in the present study was collected from 5 primary health centres in DKI Jakarta, Indonesia. Parents with child under five years old were eligible for this study. A total of 145 women fulfilled the inclusion and exclusion criteria and were analysed.

The characteristics of the parents in this study are shown in Table 4.1. The mean age of the mothers and fathers in this study was 31.06 ± 6.246 and 34.28 ± 6.782 years, respectively. The parental educational background ranged from not attending any formal school to Master’s degree graduate. Majority of mothers (57.9%) and fathers (65.5%) had senior high school education on the basis of educational status. Most mothers were unemployed (85.5%). In contrast to high unemployment rate in maternal group, majority of fathers were employed (88.3%) with 44.8% of them receiving a monthly income greater than provincial minimum wage of Rp3.940.973.

**Table 4.1.** Demographic distribution of the parents in the study.

| Variable | Total (n = 145) |  |  |  |
| --- | --- | --- | --- | --- |
|  | Maternal |  | Paternal |  |
|  | Number | % | Number | % |
| Age, years (Mean ± SD) | 31.06 ± 6.246 | | 34.28 ± 6.782 | |
| <20 | 1 | 0.7 | 0 | 0 |
| 20-24 | 19 | 13.1 | 8 | 5.5 |
| 25-29 | 42 | 29 | 38 | 26.2 |
| 30-34 | 43 | 29.7 | 37 | 25.5 |
| 35-39 | 23 | 15.9 | 30 | 20.7 |
| >39 | 17 | 11.7 | 32 | 22.1 |
| Educational background |  |  |  |  |
| Uneducated | 0 | 0 | 1 | 0.7 |
| Elementary school graduate | 9 | 6.2 | 8 | 5.5 |
| Junior high school graduate | 17 | 11.7 | 10 | 6.9 |
| Senior high school graduate | 84 | 57.9 | 95 | 65.5 |
| Diploma graduate | 10 | 6.9 | 14 | 9.7 |
| Bachelor's degree graduate | 25 | 17.2 | 16 | 11 |
| Master's degree graduate | 0 | 0 | 1 | 0.7 |
| Occupation |  |  |  |  |
| Unemployed | 124 | 85.5 | 17 | 11.7 |
| Employed | 21 | 14.5 | 128 | 88.3 |
| Household income, Rupiah |  |  |  |  |
| ≤ 500.000 | 105 | 72.4 | 4 | 2.8 |
| 500.000 -2.000.000 | 11 | 7.6 | 29 | 20 |
| 2.000.000 - 3.940.973 | 10 | 6.9 | 47 | 32.4 |
| ≥ 3.940.973 | 19 | 13.1 | 65 | 44.8 |
| House crowding index |  |  |  |  |
| Not crowded | 34 | 23.4 |  |  |
| Crowded | 32 | 22.1 |  |  |
| Severely crowded | 79 | 54.5 |  |  |
| Maternal age at delivery, year |  |  |  |  |
| <20 | 8 | 5.5 |  |  |
| 20-24 | 26 | 17.9 |  |  |
| 25-29 | 50 | 34.5 |  |  |
| 30-34 | 32 | 22.1 |  |  |
| 35-39 | 19 | 13.1 |  |  |
| >39 | 10 | 6.9 |  |  |
| Types of pregnancy |  |  |  |  |
| Singleton | 143 | 98.6 |  |  |
| Twins | 1 | 0.7 |  |  |
| Triplets | 1 | 0.7 |  |  |
| Delivery method |  |  |  |  |
| Normal delivery | 85 | 58.6 |  |  |
| Caesarean delivery | 60 | 41.4 |  |  |
| Antenatal pregnancy check-up, times |  |  |  |  |
| <5 | 10 | 6.9 |  |  |
| 5-8 | 18 | 12.4 |  |  |
| >8 | 117 | 80.7 |  |  |
| Nutrients during pregnancy |  |  |  |  |
| Adequate | 137 | 94.5 |  |  |
| Inadequate | 8 | 5.5 |  |  |
| Varied | 137 | 94.5 |  |  |
| Not varied | 8 | 5.5 |  |  |
| Eating frequency during pregnancy |  |  |  |  |
| <3 times | 6 | 4.1 |  |  |
| ≥3 times | 139 | 95.9 |  |  |
| Total pregnancy weight gain, kg |  |  |  |  |
| <6 | 21 | 14.5 |  |  |
| 6-11.5 | 71 | 49 |  |  |
| >11.5 | 53 | 36.6 |  |  |
| Stress during pregnancy |  |  |  |  |
| Yes | 38 | 26.2 |  |  |
| No | 107 | 73.8 |  |  |
| Alcohol consumption during pregnancy |  |  |  |  |
| Yes | 5 | 3.4 |  |  |
| No | 140 | 96.6 |  |  |
| Sleep habit during pregnancy, hour/day |  |  |  |  |
| <6 | 25 | 17.2 |  |  |
| ≥6 | 120 | 82.8 |  |  |
| History of complication in previous pregnancy |  |  |  |  |
| Stillbirth | 14 | 9.7 |  |  |
| Premature | 3 | 2.1 |  |  |
| Preeclampsia | 4 | 2.8 |  |  |
| Low birth weight | 5 | 3.4 |  |  |
| Medical risk during pregnancy |  |  |  |  |
| Preeclampsia | 4 | 2.8 |  |  |
| Diabetes mellitus | 2 | 1.4 |  |  |
| Smoking status |  |  |  |  |
| Active smoker | 5 | 3.4 | 80 | 55.2 |
| Ex-smoker (ceases during pregnancy) | 5 | 3.4 | 9 | 6.2 |
| Ex-smoker (ceases before pregnancy) | 3 | 2.1 | 5 | 3.4 |
| Non-smoker | 132 | 91 | 51 | 35.2 |

According to the measurement of house crowding index, majority of subjects in this study lived in severely crowded housing (54.5%). Concerning maternal age at delivery, most mothers gave birth to their children at the age of 25-29 (34.5%). Majority of the pregnancy were singleton (98.6%), while both twins and triplets independently accounted for 0.7% of the study. Vaginal birth (58.6%) dominated this study, whereas 41.4% of mothers gave birth through C-section delivery. Of all subjects, 80.7% of mothers attended more than 8 antenatal check-up visits throughout pregnancy. Food intake was adequate and varied during pregnancy in 94.5% of the mothers. About 95.9% of mothers reported eating at least 3 times per day.

Majority of the mothers experienced weight gain during pregnancy of 6-11.5 kg (49%), while about 14.5% gained less than 6 kg and about 36.6% gained more than 11.5 kg. In this study, 26.2% percent of mothers reported experiencing stress throughout pregnancy, while alcohol consumption was observed in 3.4% mothers. About four-fifth of the mothers reported sleeping for at least 6 hours daily. History of pregnancy complication was observed in 17.9% of mothers, with stillbirth being the most common complication. Preeclampsia was observed in 2.8% of the sample population, while diabetes mellitus during pregnancy was observed in 1.4% of the subjects. Of all parents, 5 mothers and 80 fathers were actively smoking. Further information on parents’ demographic characteristics is provided in Table 4.1.

Five mothers and eighty fathers were actively smoking, so we collected information regarding their smoking habit, including type of cigarettes, duration of smoking, number of cigarettes, and location of smoking. Of 5 mothers who were active smokers, all smoked using cigarette sticks (100%), and 3 out of 5 smoked for more than 1 hour (60%). Three mothers were smokers of 1-10 cigarettes per day (60%), while two mothers smoked 11-20 cigarettes daily (40%). Two mothers reported smoking at home (40%), whereas the remaining three smoked at places other than home (60%). From paternal side, about 98.8% of fathers used cigarette sticks, while 1.3% used electronic cigarettes. The percentage of fathers smoking for less or equal to an hour (56.3%) was slightly higher compared to those who smoked for more than 1 hour (43.8%). Majority of fathers smoked 1-10 cigarette sticks per day (73.8%), while smokers of 11-20 and 21- 30 cigarettes accounted for 18.8% and 7.5% of all actively smoking fathers, respectively. About 65% of fathers reported smoking at places other than home, demonstrating greater percentage as compared to those who smoked at home (35%). The summary of actively smoking parents’ characteristics is depicted in Table 4.2.

**Table 4.2.**
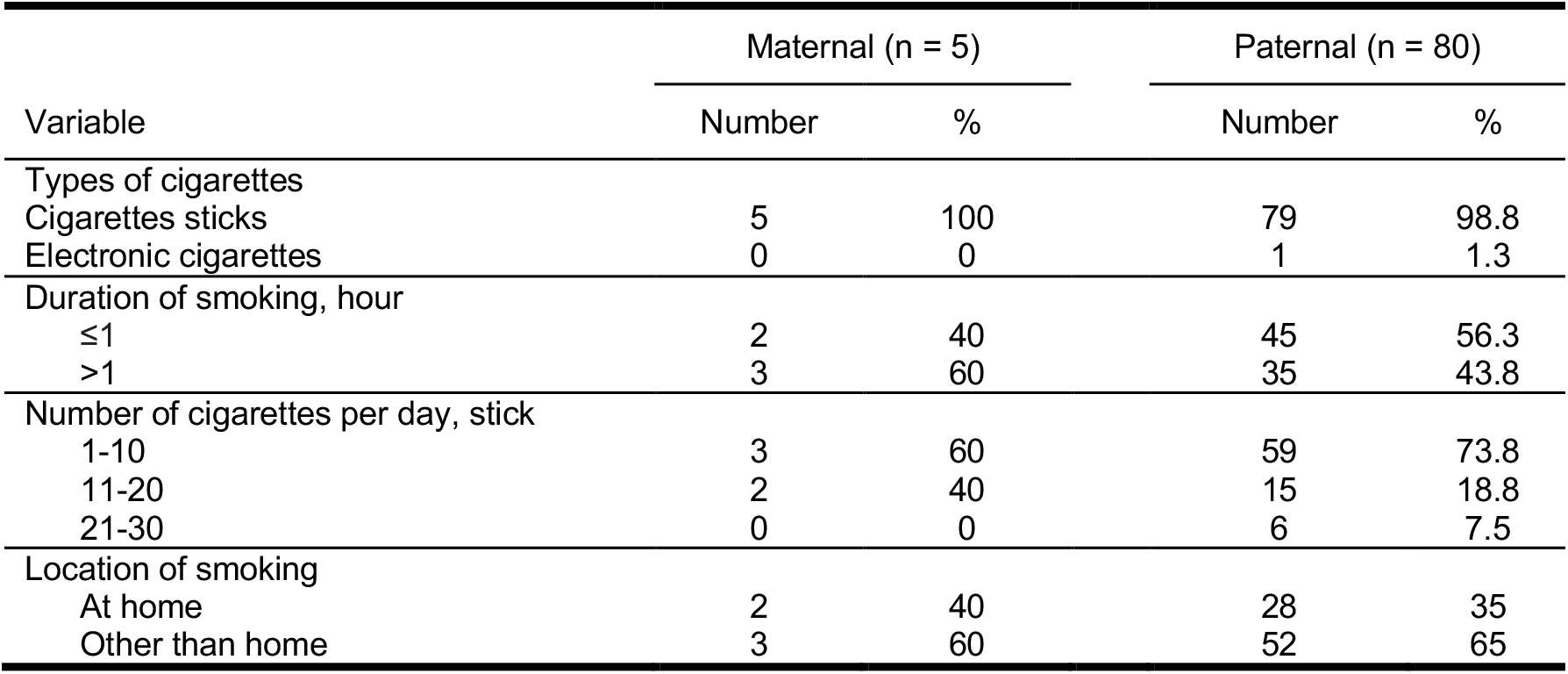
Characteristics of actively smoking parents in the study.

Table 4.3 depicts the information on children’s demographic characteristics in the study. More female (57.9%) than male (42.1%) children were observed in this study. Majority of the children were their mothers’ first-borns (60%). The mean birth weight of children was 3.117 ± 0.515 kg. Of 145 subjects, 16 of the children were categorized as low birth weight (<2500 grams), while the remaining 129 children’s birth weight were considered normal. Majority of the children were born at early term (61.4%) and the mean of gestational age was 37.366 ± 1.874 weeks.

**Table 4.3.** Demographic distribution of the children in the study.

| Variable | Total |  | Mean ± SD |
| --- | --- | --- | --- |
|  | Number | % |  |
| Gender |  |  |  |
| Male | 61 | 42.1 |  |
| Female | 84 | 57.9 |  |
| Birth order |  |  |  |
| First born | 60 | 41.4 |  |
| Second born | 50 | 34.5 |  |
| Third born | 22 | 15.2 |  |
| Fourth born | 10 | 6.9 |  |
| Fifth born | 3 | 2.13 |  |
| Birth weight, g |  |  |  |
| <2500 (low birth weight) | 16 | 11 | $3.117 \pm 0.515$ |
| ≥2500 (normal birth weight) | 129 | 89 |  |
| Gestational age, week |  |  |  |
| 28-31 (very preterm) | 3 | 2.1 | $37.366 \pm 1.874$ |
| 32-36 (moderate/late preterm) | 23 | 15.9 |  |
| 37-38 (early term) | 89 | 61.4 |  |
| 39-40 (full term) | 28 | 19.3 |  |
| 41 (late term) | 2 | 1.4 |  |

### Proportions of Low Birth Weight Infants Across Categories of Paternal and Maternal Smoking

There was a dose-response relationship between paternal number of cigarettes/day and duration of smoking with low birth weight. In fathers who smoked 11-30 cigarettes/day, 23.81% of the offspring were born with low birth weight, which is higher compared to those that smoked 1-10 cigarettes/day (13.56%) and did not smoke throughout pregnancy (4.61%). Similarly, the prevalence of low birth weight was higher in fathers who smoked for more than an hour (17.14%) than those who smoked for less than an hour (15.5%) and did not smoke at all (4.61%). This showed that fathers smoking more cigarettes and for longer period of time were related with higher prevalence of low birth weight in this study. In contrast, there was no linear interaction between maternal smoking and low birth weight, which might be primarily due to a fairly small number of actively smoking mothers (3.4%) in the present study.

### Bivariate Analysis of the Association Between Parents’ and Children’s Demographic Characteristics and Birth Weight

This study identified the association between several factors, including maternal age, maternal educational level, socio-economical background, medical risks, environmental and behaviour risks, as well as paternal smoking habit. To identify possible confounders to control for in the assessment, a bivariate analysis was done, with its results presented in Table 4.4. Based on Pearson chi-square value, birth order (p = 0.018) and paternal smoking status (p = 0.026) were significant predictor of low birth weight in this study due to its p value of <0.05. First-borns were found to be 3.592 times more likely to be born with low birth weight compared to second to fifth child. Tobacco exposure from actively smoking fathers during pregnancy led to 4.010 times higher risk for the occurrence of low birth weight compared to no paternal tobacco exposure. Fisher’s exact test is used to measure significance in variables showing variable value less than 5. From Fisher’s exact test, maternal educational background (p = 0.042), gestational age (p = 0.002), maternal age at delivery (p = 0.004), nutrients during pregnancy (p = 0.044), and history of pregnancy complication (p = 0.042) showed significant association with low birth weight. Other than these factors, the remaining factors were found to be not significantly associated with the occurrence of low birth weight.

**Table 4.4.**
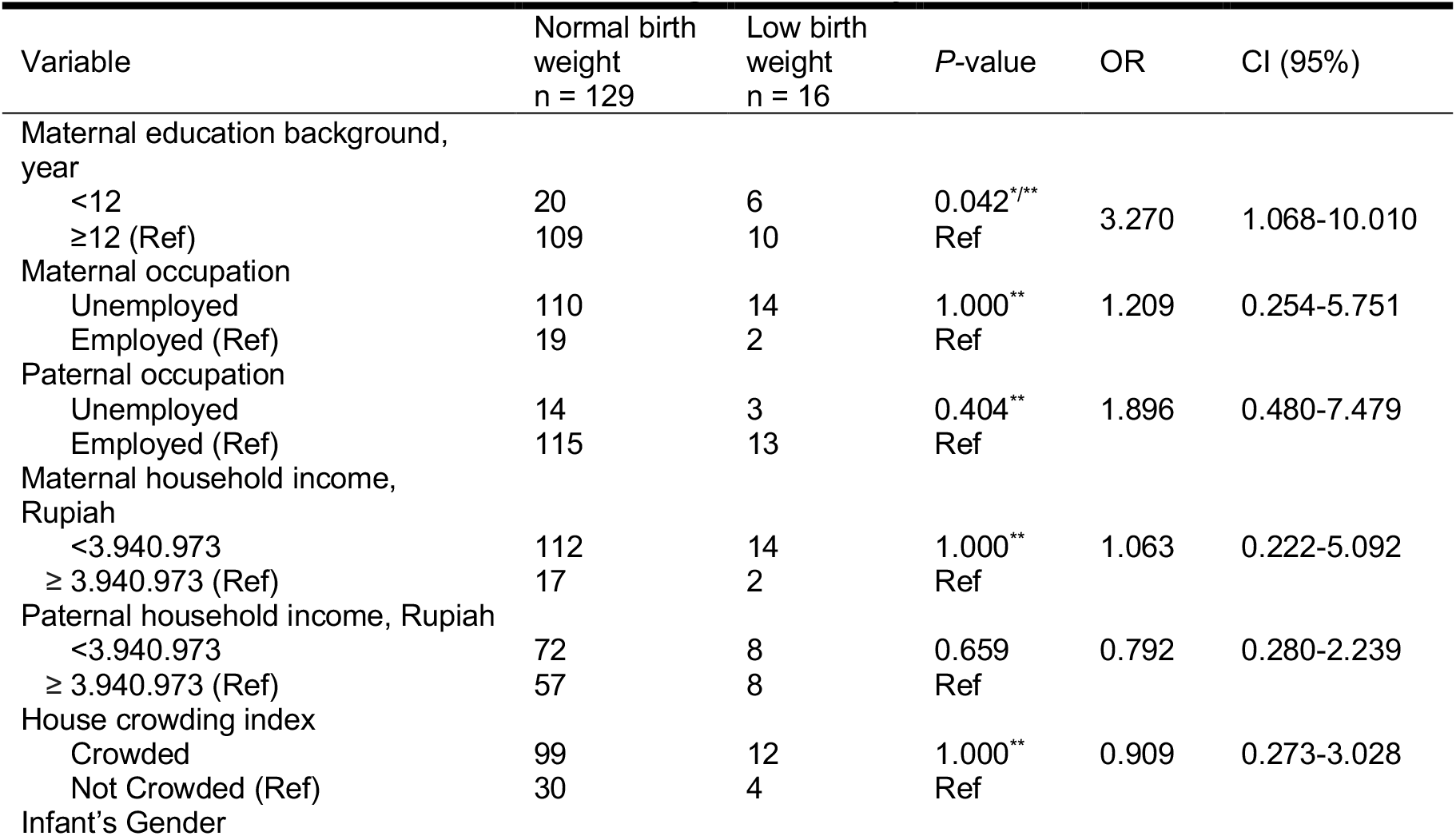
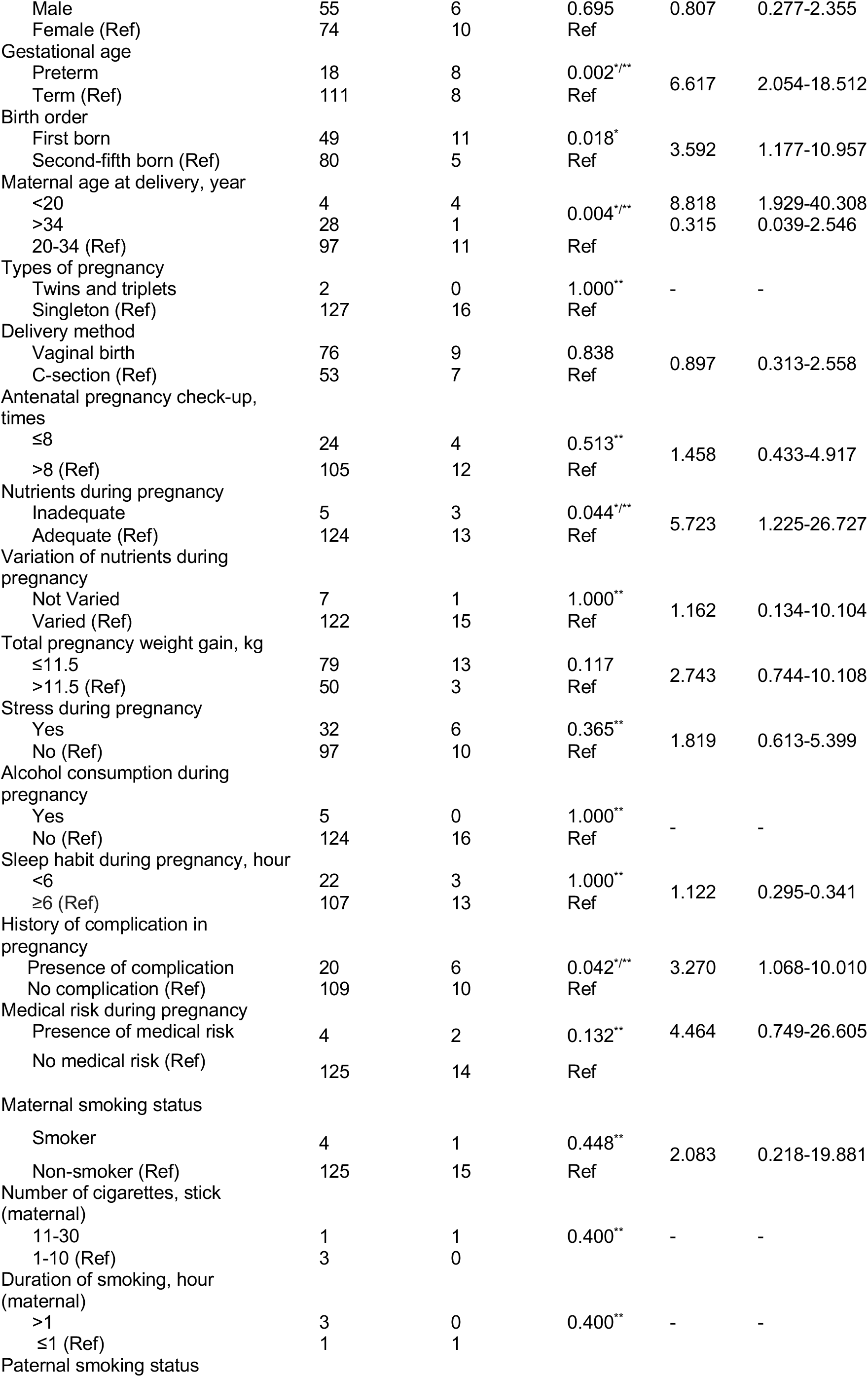
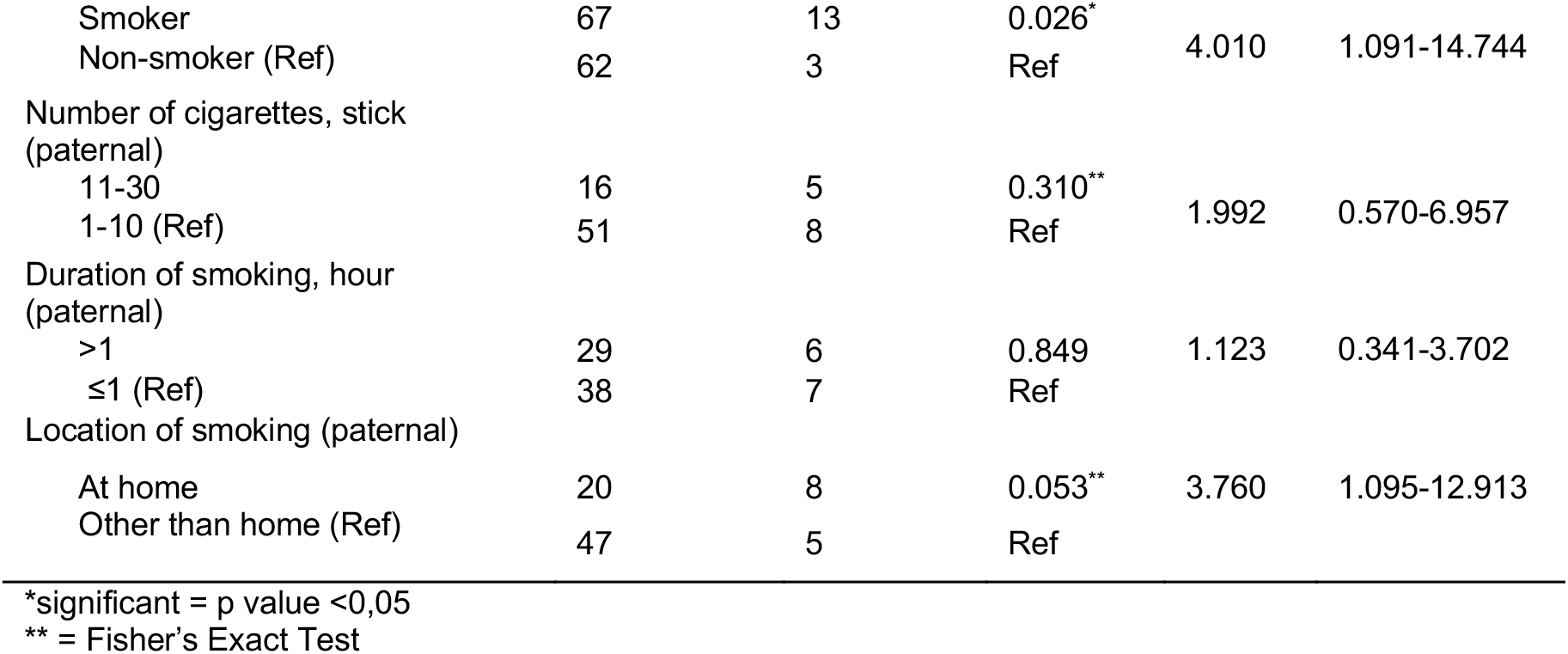
Factors associated with low birth weight in the study.

### Multivariate Logistic Regression Analysis

To control for interrelations between different variables, a multivariate logistic regression analysis was done. The logistic regression was performed to test the effects of paternal educational background, maternal educational background, gestational age, birth order, maternal age at delivery, nutrients during pregnancy, total pregnancy weight gain, history of pregnancy complication, presence of medical risk during pregnancy, paternal smoking status, and paternal location of smoking on the occurrence of low birth weight.

In this analysis, the dependent variable is coded into 1 and 0. “The occurrence of low birth weight” is referred as code 1, while “no low birth weight” is given the code 0 as the reference category in the analysis. From omnibus test of model coefficients, the logistic regression model was statistically significant, χ^2^(10) = 42.476, p = 0.000 (<0.05). Since the significance value is less than 0.05, therefore the null hypothesis (H0) is rejected, indicating that the current model outperforms the null model. The R^2^ value describes how much variation in the outcome is approximately explained by the model. The Nagelkerke R^2^ indicates that the model accounted for 50.7% of the total variance.

The Hosmer and Lemeshow test of the goodness of fit demonstrates that the model is a good fit to the data and can be approved as p = 0.286 (>0.05). From the classification table, the sensitivity of prediction is 98.4%, while the specificity of prediction is 62.5%. The overall predictive accuracy of this research model is 94.5%.

Attendance of <12 years of school from maternal side; preterm gestational age; first-borns; maternal age at delivery <20 years; maternal age at delivery >34 years; inadequate food intake during pregnancy; weight gain of ≤11.5 kg during pregnancy; history of pregnancy complication; presence of pregnancy medical; and tobacco exposure from paternal side were all identified as possible confounders (p < 0.25). The association between these factors and occurrence of low birth weight, adjusted for interrelations between different variables, is shown in Table 4.5.

**Table 4.5.** Multivariate Logistic Regression Analysis.

| Variable | B | SE | P value | OR | 95% CI for OR |  |
| --- | --- | --- | --- | --- | --- | --- |
|  |  |  |  |  | Lower | Upper |
| Mothers attending <12 years of school | 1.009 | 0.768 | 0.189 | 2.743 | 0.609 | 12.363 |
| Preterm gestational age | 1.737 | 0.751 | 0.021 | 5.683 | 1.305 | 24.753 |
| First-borns | 1.756 | 0.881 | 0.046 | 5.787 | 1.030 | 32.520 |
| Age at delivery: <20 years | 1.382 | 1.025 | 0.178 | 3.982 | 0.534 | 29.701 |
| Age at delivery: >34 years | -1.998 | 1.498 | 0.182 | 0.136 | 0.007 | 2.554 |
| Inadequate food intake during pregnancy | 2.854 | 1.314 | 0.030 | 17.364 | 1.322 | 228.051 |
| ≤11.5 kg weight gain during pregnancy | 1.134 | 0.827 | 0.171 | 3.107 | 0.614 | 15.720 |
| History of pregnancy complication | 1.367 | 0.880 | 0.120 | 3.922 | 0.699 | 22.015 |
| Presence of pregnancy medical risk | 0.549 | 1.931 | 0.776 | 1.732 | 0.039 | 76.244 |
| Actively smoking fathers | 2.176 | 0.960 | 0.023 | 8.807 | 1.341 | 57.833 |
| Constant | -6.749 | 1.440 | 0.000 | 0.001 |  |  |

In multivariate analysis, positive and statistically significant association were found between positive preterm gestational age (β = 1.737, p = 0.021), first-born children (β = 1.756, p = 0.046), inadequate food intake during pregnancy (β = 2.854, p = 0.030), actively smoking fathers (β = 2.176, p = 0.023) and occurrence of low birth weight. Preterm infants were 5.683 times more likely to be born with low birth weight compared to infants born with term gestational age. First-borns in family were at 5.787 times higher risk to be born with LBW. Mothers who did not consume adequate food during pregnancy were found to be 17.364 times more likely to give birth to low birth weight children compared to those consuming adequate food intake. Actively smoking fathers increased the risk of low birth weight by 8.807 times. No statistically significant association was found between mothers attending <12 years of school (p = 0.189), age at delivery <20 years (p = 0.178), age at delivery >34 years (p = 0.182), weight gain of ≤11.5 kg during pregnancy (p = 0.171), history of pregnancy complication (p = 0.120), presence of pregnancy medical risk (p = 0.776), and occurrence of low birth weight in infants.

## Discussion

Low birth weight (LBW) is a major determinant of infant morbidity and mortality. It is a major public health problem due to its association with infant survival and risk of developmental disabilities and illnesses. In infants weighing 1500-2500 grams, it is estimated that the risk of neonatal death significantly rises to 20 times higher compared to those of normal birthweight. More significantly, the risk of death increases to 100 times in infants born with very low birth weight (VLBW). Globally, the prevalence of LBW is about 15.5% of all births, reflecting 20 million infants born with LBW each year.^13^ This study investigated the association between tobacco exposure during pregnancy and other risk factors and occurrence of LBW.

### General Characteristics of Children in Study Population

Of 145 subjects, there were more female (57.9%) compared to male children participating in this study. Most children were first-borns (41.4%) and had normal birth weight (89%). About 18% children were born prematurely and the remaining 82% were born full-term. The average birth weight and gestational age in this study were 3.117 ± 0.515 kg and 37.366 ± 1.874, respectively. According to Indonesian Children Profile in 2019, there are more male compared to female (1.02:1) children aged below 1 year in Indonesia. The data also revealed that about 82.14% of pregnant women in Indonesia gave birth to children with normal birth weight.^14^ Preterm birth rate in Indonesia is 15 per 100 births, placing Indonesia at the fifth place in the world for the number of premature births.^15^

### Prevalence of Low Birth Weight

In our study, the prevalence of term LBW was 11%, which is slightly below global prevalence of LBW in developing countries of 15.5%.^13^ Our study demonstrated that sex of the child was not associated with LBW. However, a study demonstrated that male sex is more vulnerable to perinatal mortality and morbidity, including IUGR, low Apgar score and respiratory insufficiency.^16^ In Indonesia, according to Indonesian Children Profile 2019, about 13% of all births were born with low birth weight in the past 2 years. The incidence was slightly higher in rural areas (13.18%) compared to in urban areas (12.84%).^14^ In other studies conducted in Indonesia, the prevalence of LBW ranged between 6.71- 10.2%.17,18,19

### Association Between Tobacco Exposure and Low Birth Weight

Our finding revealed that tobacco exposure from paternal side during pregnancy was positively associated with birth weight of the newborns (p = 0.023, adjusted OR = 8.794, 95% CI = 1.344- 57.540). Of 16 LBW infants in the study, 13 infants (81.25%) were exposed to passive tobacco exposure from their fathers, whereas 18.75% of LBW infants were infants of unexposed mothers. Therefore, in our study, the prevalence of LBW among infants of secondhand smoke-exposed mothers and unexposed mother was 16.25% and 4.6% respectively. This study assesses the number of cigarettes smoked, duration and location of tobacco exposure. It was found that higher amount of cigarettes smoked (p = 0.310) and higher duration of tobacco exposure (p = 0.849) were not significantly associated with LBW. Location of smoking, although not statistically significant, might act as a confounding factor (p = 0.053, crude OR = 3.760, 95% CI = 1.095-12.913), demonstrating that fathers who were smoking at home during pregnancy might put the mothers at higher risk of giving birth to LBW children compared to those smoking at other places. After interrelations between different variables were adjusted, paternal smoking status was positively and significantly associated with lower birthweight, while location of smoking was not a statistically significant predictor.

These findings were supported by previous studies showing that children born to smoking fathers had a significant reduction in birthweight and increased risk of LBW.^20,21^ A research in Denmark demonstrated that passive tobacco exposure from fathers was significantly associated with a reduction of birth weight of 120 gram per pack of cigarettes smoked per day.^22^ Although our study did not reveal significant association from number of cigarettes smoked and duration of smoking, prior studies found a significant dose-effect relationship between number of cigarettes smoked per day and new- borns’ birth weight.^22,23^ Similarly, a study found that paternal smoking amount of >20 cigarettes/day (OR = 2.09, 95% CI = 1.38-3.17, p = < 0.001) put the infants at a significantly higher risk of having LBW compared to fathers who smoked 11-20 cigarettes/day (OR = 0.66, 95% CI = 0.46- 0.94, p = 0.022) and 1-10 cigarettes/day (OR = 0.81, 95% CI = 0.58-1.14, p = 0.230).^20^ A study conducted among Indonesian mothers revealed that duration of exposure per day of more than 60 minutes resulted in higher incidence of LBW (91.7%) compared to exposure of 15-60 minutes (48.1%) and less than 15 minutes per day (34.5%) (p = 0.004).^24^

From maternal side, there was no significant association between newborns’ birth weight and firsthand smoke exposure from actively smoking mothers during pregnancy (p = 0.448, OR = 2.083, 95% CI – 0.218-19.881). Similarly, our study revealed that maternal number of cigarettes smoked (p = 0.400) and duration of firsthand smoke exposure (p = 0.400) had no significant association with LBW, probably reflecting the small number of actively smoking mothers in the study (3.45%). Of 16 LBW infants in the study, only 1 infants (6.25%) was exposed to first-hand tobacco exposure from the mother. Previous studies demonstrated that maternal smoking did not show statistically significant association with increased risk of LBW, but rather a reduction in birth weight.^25,26^ However, several prior studies contradict with this finding, suggesting significant association of LBW in relation to active maternal tobacco exposure.^27,28,29^ Several studies reported that the most significant effect of maternal smoking on birth weight occurs during late pregnancy, particularly in heavy smokers (>8-10 cigarettes per day). Moreover, infants of mothers who smoked >20 cigarettes/day was most susceptible to deliver LBW offspring, irrespective of stage of pregnancy.^27,28^ In third trimester, a 27-g reduction of birth weight was found in each cigarette smoked per day.^27^ Changing smoking habit is found to be most beneficial if the mother quits smoking in early pregnancy. This is supported by a study in Netherlands demonstrating that pregnant women who quitted smoking after recognizing their pregnancy had a decreased risk of LBW compared to those who continued smoking or reduced the amount of cigarettes smoked without quitting completely.^28^

A crucial consideration in this study is that the validity and accuracy of the number of cigarettes, duration of tobacco exposure, and location of smoking might limit the credibility of this study.

### Role of Tobacco in Low Birth Weight Newborns

The observed association between tobacco exposure during pregnancy and birth weight of newborns might indicate several biological mechanisms directly or indirectly leading to sub-par development of the fetus. In general, nicotine, a principal alkaloid of tobacco cigarette, gains access to the fetal compartment through the placenta. Fetal concentration is generally 15% higher compared to maternal serum level. The mean ratio between placental tissue/maternal serum was 2.58 ± 1.3. As nicotine is absorbed orally, *in utero* swallowing exposes fetus to nicotine. Therefore, exposure to tobacco during pregnancy leads to significant fetal exposure to nicotine during intrauterine life.^30^

Nicotine has been found to mediate constriction of intrauterine vessels and lead to increased placental syncytiotrophoblast proliferation. In smokers, potentially harmful deoxyribonucleic acid (DNA) adducts (metabolic products of polycyclic aromatic hydrocarbons [PAH]) are found to cross or collect in the placenta. Nitrosamines and PAH together act as carcinogenic species in tobacco, which are metabolized in a series of 2-phase enzymatic metabolic reactions. The first phase consists of metabolic activation of PAH compounds by CYP1A1 enzymes. This reaction results in reactive oxygen intermediates that are able to covalently bind DNA to form adducts.^31^

The second phase metabolic reaction aims to detoxify these reactive electrophilic intermediates with the help of glutathione S-transferase (GSTT1). This reaction, through conjugation with endogenous species, forms hydrophilic glutathione conjugates that are readily excreted. In pregnancies exposed to tobacco, several studies demonstrated that deletion of fetal GSST1 and increased placental CYP1A1 might be significantly associated with neonatal birthweight reduction. Thus, the extend of the damage and adverse outcomes might be determined by the coordinated expression as well as the relative balance of these enzymes. Although the exact underlying mechanisms resulting in growth restriction following *in utero* tobacco exposure remain poorly understood, they are frequently attributed to chronic fetal hypoxia due to vasoconstriction of uteroplacental blood vessels. This results in reduced placental blood flow and reduction in oxygen and nutrient delivery to the fetus.^31^

### Prevention and Management

LBW is regarded as an adverse pregnancy outcome that is associated with many risk factors either during pre-pregnancy or gestation, or both. It is recognized that the etiology of LBW is multifactorial and might vary by setting.^32^ In the present study, we have shown that LBW was significantly associated with paternal tobacco exposure during pregnancy, premature delivery, first birth order and inadequate food intake during pregnancy.

Many risk factors can be identified before pregnancy occurs. In our study, about 5.5% pregnant women and 9.6% of their partners quit smoking before or during pregnancy. Cessation of smoking happens mainly due to increased awareness about the adverse pregnancy outcome and long-term post-natal health effects on the offspring. According to a study conducted in 2016, participants in their study acknowledged that smoking during pregnancy could be harmful and resulted in adverse outcome. However, despite knowing the harmful effects, many continued to smoke during pregnancy and only a few held negative attitudes and beliefs regarding this behaviour. Tackling tobacco cessation is indeed challenging. This study highlighted the importance of public health interventions in raising the awareness of adverse outcomes of tobacco exposure to increase women’s intention to quit smoking, either before or during pregnancy. This study also encouraged health care providers to incorporate conversations regarding the smoking cessation during pregnancy as well as the importance of quitting completely among reproductive-aged women.^33^

### Research Strength

This is the first cross-sectional study conducted to analyse the association between maternal and paternal tobacco exposure during pregnancy with prevalence of LBW at a population level in DKI Jakarta.

### Research Limitation

Several limitations of this study need to be addressed. First, some confounding factors were not scrutinized. BMI of mothers before pregnancy, interval of pregnancy, maternal HIV infection, and other environmental factors could potentially correlate to LBW and confound the relation of tobacco exposure to birth weight. Second, information regarding smoking habits of parents in this study was collected by self-report and there might be misclassification between smoking and non-smoking group. Third, it should be noticed that the number of sample is fairly smaller compared to other research. Fourth, the available data were collected at the time of distributing the questionnaire, and not during pregnancy. Several variables might differ between the time of pregnancy and time of survey. Lastly, only 3.4% of the mothers in the study smoked which might lead to reduced statistical power.

## Conclusion

In our cross-sectional study, the proportion of LBW was 11%. Of 16 LBW infants, 1 infant (6.25%) was exposed to first-hand maternal tobacco exposure and 13 infants (81.25%) were exposed to second-hand paternal tobacco exposure during pregnancy. Our study demonstrated that second-hand tobacco exposure from actively smoking fathers is responsible for significantly increased prevalence of low birth weight. In contrast, the association between first-hand maternal tobacco exposure and low birth weight is insignificant. Although not statistically significant, there was a dose-response relationship between paternal number of cigarettes/day and duration of smoking with LBW. However, both paternal and maternal number of cigarettes/day and duration of smoking are not significant predictors of LBW. Multivariate logistic regression analysis in the present study showed that premature delivery, birth order and inadequate food intake during pregnancy are significant predictors of low birth weight.

## Data Availability

Due to the nature of this research, participants of this study did not agree for their data to be shared publicly, so supporting data is not available.

## Acknowledgement

Authors must not fail to appreciate the Ethics Committee of Faculty of Medicine Universitas Indonesia for Hibah PUTI 2020 that helped us in funding this research

